# Urinary cell mRNA profiling of kidney allograft recipients: Development of a portable protocol for noninvasive diagnosis of T cell mediated rejection and BK virus nephropathy

**DOI:** 10.1101/2022.09.15.22279980

**Authors:** Thalia Salinas, Carol Li, Catherine Snopkowski, Gabriel Stryjniak, Divya Shankaranarayanan, Shady Albakry, Ruchuang Ding, Vijay K. Sharma, Steven P. Salvatore, Surya V. Seshan, Darshana M. Dadhania, Thangamani Muthukumar, Manikkam Suthanthiran

## Abstract

**Background:** We developed urinary cell mRNA profiling for the noninvasive diagnosis of acute T cell mediated rejection (TCMR) and BK virus nephropathy (BKVN), two significant post-transplant complications. Our profiling protocol for the multicenter Clinical Trial of Transplantation-04 (CTOT-04) study consisted of centrifugation of urine to prepare cell pellets, washes, addition of an RNA preservative, storage at -80^0^ C and shipment in cold containers to our Gene Expression Monitoring Core for total RNA isolation and quantification of mRNA copies in RT-qPCR assays. To simplify profiling, we developed a filter-based protocol (ZFBP) that eliminated the need for centrifugation, RNA preservative, storage at -80^0^ C, and shipment in cold containers for mRNA profiling; furthermore, kidney allograft recipients could be trained to perform the filtration of urine at home using the filter and post the urinary cell lysate containing the total RNA at ambient temperature to our Core for profiling. We have now refined ZFBP and investigated the diagnostic performance characteristics of a protocol designated as Weill Cornell Hybrid Protocol (WCHP).

**Methods:** Total RNA was isolated from kidney allograft biopsy matched urines from kidney allograft recipients using a filter-based protocol complemented by a silica-membrane based cartridge for mRNA enrichment (WCHP). Absolute copy numbers of CD3ε mRNA, CXCL10 mRNA and 18S ribosomal RNA, components of the CTOT-04 three gene TCMR diagnostic signature, and urinary cell BKV VP 1 mRNA copy number were measured using RT-qPCR assays. Mann-Whitney test, Fischer exact test and receiver operating characteristic (ROC) curve analysis were used for data analyses.

**Results:** Urinary cell three gene TCMR diagnostic signature in urines processed using the WCHP discriminated kidney allograft recipients with TCMR (n=12 biopsies from 11 patients) from those without TCMR (n=29 biopsies from 29 patients). The median (25^th^ and 75^th^) score of the CTOT-04 three gene TCMR diagnostic signature was -0.448 (-1.664, 0.204) in the TCMR group and -2.542 (-3.267, -1.365) in the No Rejection biopsy group (P=0.0005, Mann-Whitney test). ROC curve analysis discriminated TCMR group from the No Rejection group and the area under the ROC curve (AUROC) was 0.84 (95% Confidence Intervals [CI], 0.69 to 0.98) (P<0.001), and TCMR was diagnosed with a sensitivity of 67% (95% CI, 35 to 89) at a specificity of 86% (95% CI, 67 to 95) using the CTOT-04 validated cutpoint of -1.213 (P=0.0016, Fischer exact test). BKV VP1 mRNA copy number in urines processed using the WCHP discriminated patients with BKVN (n=7) from those without BKVN (n=29) and the AUROC was 1.0 (95% CI, 1.00 to 1.00) (P<0.0001) and BKVN was diagnosed with a sensitivity of 86% (95% CI, 42 to 99) at a specificity of 100% (95% CI, 85 to 100) with the previously validated cutpoint of 6.5 × 10^8^ BKV-VP1 mRNA copies per microgram of total RNA (P<0.0001, Fischer exact test).

**Conclusion:** Urine from kidney allograft recipients processed using the WCHP predicted TCMR and BKVN. WCHP represents not only a significant advance towards portability of urinary cell mRNA profiling but also improved patient management by minimizing their visits for urine collection.

## 1. Introduction

Kidney transplantation is the treatment of choice for patients with end stage kidney disease (Wolfe et.al., 1999; Hart et al., 2020). The realization of the full benefits of transplantation however is undermined by immune rejection of the kidney allograft, and acute T cell mediated rejection (TCMR) is the most frequent type of immune rejection (Suthanthiran et al., 1994; Rampersad et al., 2022). Infection is another threat for the continued functioning of the transplanted kidney, and BK virus nephropathy (BKVN) has emerged as a serious and frequent post-transplant complication (Bohl et al., 2007; Hirsch et al., 2013).

Percutaneous needle core biopsy of the kidney allograft is the standard-of-care for the diagnosis of both TCMR and BKVN, but the invasive biopsy procedure is associated with multiple complications including bleeding, graft loss and even death, albeit in rare circumstances (Morgan et al., 2016; Redfield et al., 2016; Plattner et al., 2018). The well documented inter-observer variability in the reading of the allograft biopsy is another valid concern (Furness et al., 2001; Veronese et al., 2005). Also, the immune response is dynamic, and it is neither safe nor practical to perform repeated biopsies to capture the kinetics of the anti-allograft repertory.

To facilitate safer interrogation of kidney allograft status, we developed and refined urinary cell mRNA profiling for the noninvasive diagnosis of TCMR (Li et al., 2001; Muthukumar et al., 2003; Ding et al., 2003; Dadhania et al., 2003; Tatapudi et al., 2004; Muthukumar et al., 2005; Afaneh et al., 2010; Anglicheau et al., 2012). Our single center study led to a multicenter Clinical Trial of Transplantation-04 (CTOT-04) study of 485 kidney allograft recipients in which we profiled 4300 urine specimens and discovered and validated a parsimonious urinary cell three gene TCMR diagnostic signature of T cell CD3ε mRNA, mRNA for the proinflammatory chemokine IP10 (CXCL10) and 18S ribosomal RNA (rRNA), a member of the transcriptional machinery (Suthanthiran et al., 2013).

In view of the clinical significance of BKVN, we also designed oligonucleotide sense and antisense primers and TaqMan probe for the absolute quantification of BKV VP1 mRNA copy number in the RT-qPCR assay for the noninvasive diagnosis of BKVN (Ding et al., 2002) and validated using an independent cohort of kidney allograft recipients that BKV VP1 mRNA copy number threshold of 6.5×10^8^ or greater per microgram of total RNA is diagnostic of BKVN in an independent cohort of kidney allograft recipients (Dadhania et al., 2010).

In the multicenter CTOT-04 study, urine samples were centrifuged to prepare urinary cell pellets at each participating academic transplant center. Centrifugation of the urine sample and the associated steps to prepare urinary cell pellets required technical expertise and laboratory equipment including different types of centrifuges and the urinary cell pellets were stored at -80^0^ C and shipped to our Gene Expression Monitoring (GEM) Core in specialized cold containers.

Centrifugation of urine samples was also the protocol in our studies of BKVN. To reduce the complexities in the initial steps of urine processing, we recently developed a Zymo ZRC GF™ filter-based protocol (ZFBP) that eliminated the need for centrifugation, the RNA preservative, storage at -80 ^0^C, and shipment of specimen on dry ice to our GEM Core. Furthermore, the kidney graft recipients could be trained to perform the initial filtration step at home and ship the urinary cell lysate containing the total RNA at ambient temperature to our GEM Core for downstream urinary cell mRNA profiling (Snopkowski et al., 2021).

In the current investigation, we retained the advantages of the ZFBP but modified the procedure to enrich for mRNA in the total RNA isolated from the urine sample. We accomplished mRNA enrichment by incorporating a spin column from the Qiagen RNeasy Mini Kit (Cat.No.74104) to isolate total RNA instead of using the Zymo-Spin™ IC column included in the ZR Urine RNA isolation Kit (Cat. No. RI038). The silica-membrane based spin column from the RNeasy mini kit excludes RNA molecules shorter than 200 nucleotides and enriches for mRNA (RNeasy Mini Handbook 2019) whereas all RNA molecules greater than 17 nucleotides are eluted using the Zymo-Spin™ IC column, an advantage for profiling small RNAs such as miRNAs. Our modified hybrid procedure, designated as Weill Cornell Hybrid Protocol (WCHP), was used in this investigation to isolate total RNA in the urinary cell lysates collected using the Zymo ZRC GF™ filter. Our objectives were to determine whether noninvasive diagnosis of TCMR with performance characteristics observed in the CTOT-04 study is feasible using the WCHP, and whether noninvasive diagnosis of BKVN with performance characteristics observed in the BKVN validation study is feasible using the WCHP.

## 2. Materials and methods

### 2.1 Study cohorts and biospecimens

Sterile urine cups were used to collect approximately 50 mL of urine from 45 unique kidney allograft recipients at the time of percutaneous needle core biopsies (biopsy-matched urine samples) of the kidney allograft. The biopsies were stained with hematoxylin and eosin, and periodic acid-Schiff and classified by Weill Cornell nephropathologists using the Banff kidney allograft biopsy classification schema (Loupy et al., 2017). PAb416 mouse monoclonal antibody (mAb) directed at the simian virus (SV) 40 large T antigen and cross reactive with the BKV large T antigen (Mann et al., 1984) is the mAb used by our center pathologists to detect intragraft presence of BK virus in clinical samples, and the biopsies included in the current study were stained with PAb416 mAb and classified as BKVN on the basis of positive intragraft staining for SV40/BKV large T antigen and histological features consistent with BKVN (Drachenberg et al., 2004). All biopsy classification were made by the nephropathologists masked to urinary cell gene expression patterns.

Kidney allograft recipients signed written informed consent for participation under one of the following protocols: IRB Protocol# 9402002786 “Use of PCR to Evaluate Renal Allograft Status”, IRB Protocol#0710009490 “Evaluation of Subclinical Renal Allograft Damage” and/or the IRB Protocol # 1207012730 “Development of Gene Expression Monitoring Bio-Bank to Study Non-Invasive Biomarkers that Diagnose and Anticipate Post-Transplant Complications.” All protocols were approved by the Weill Cornell Medicine Institutional Review Board. The research study was conducted as per the ethical principles outlined in the Declaration of Helsinki (Puri et al., 2009).

### 2.2. Isolation of RNA from urine specimens

We isolated total RNA from urine specimens using the novel WCHP. The WCHP protocol differed from the previously described Zymo filter based protocol (Gomez-Alamillo et al., 2010; Bradley et al., 2019; Snopkowski et al., 2021) in that the total RNA in the urinary cell lysates collected after the urine is pushed through the ZRC GF™ filter is isolated using the RNeasy Mini Kit (Qiagen, Cat. No. 74104) and not with the RI038 ZR Urine RNA Isolation Kit (ZYMO, Cat. No. R1038). In the CTOT-04 study, PureLink Micro-to-Midi total RNA purification system (Cat. No. 12183018, Invitrogen) was used to isolate total RNA from the urine. A side-by-side comparison of the RNeasy Mini Kit and PureLink RNA Mini Kit showed that both kits yield high-quality RNA as determined by spectrophotometry, agarose gel, and 2100 Bioanalyzer, and had similar quantities of residual DNA (Invitogen). We measured total RNA yield (absorbance at A260) and its purity (ratio of A260/A280) using the Nanodrop One Spectrophotometer (ThermoFisher Scientific).

### 2.3. Reverse transcription of total RNA to complementary cDNA

Total RNA was reverse transcribed to cDNA using the TaqMan reverse transcription kit (Applied Biosystems, Cat. No. N808-0234). Total RNA, at a concentration of 1.0 μg in 100 μl volume, was reverse transcribed and the reverse transcription reaction contained 1x TaqMan reverse transcription buffer, 500 μM each of 4 dNTPs, 2.5 μM of Random Hexamer, 0.4 Unit/μl of RNase inhibitor, 1.25 Unit/μl of MultiScribe Reverse Transcriptase and 5.5 mM of Magnesium Chloride. The RT mixture was incubated at 25 °C for 10 min, 48 °C for 30 min, and 95 °C for 5 min.

### 2.4. Design of Gene Specific Oligonucleotide Primers and TaqMan Probes

We designed gene specific oligonucleotide primers and TaqMan (hydrolysis) fluorogenic probes for the absolute quantification CD3ε mRNA, CXCL10 mRNA, TGFβ1 mRNA and 18S rRNA in RT-qPCR assays. We have developed and reported the sequence and location in the genome of the gene specific oligonucleotide primers and of TaqMan probes for the quantification of CXCL10 mRNA, TGFβ1 mRNA and 18S rRNA in RT-qPCR assays (Suthanthiran et al., 2013). For the absolute quantification of CD3ε mRNA, we used the previously reported gene specific oligonucleotide primers in combination with an improved design of the TaqMan probe. In the current study, major groove binding protein (MGB) replaced TAMRA and the following probe we designed was used for the quantification of CD3ε mRNA: 5’ FAM TCTGGAACCACAGTAATA MGB 3’. The replacement of TAMRA dye in the TaqMan reporter with MGB eliminates the background fluorescence of the quencher TAMRA and allows for the design of a shorter probe with increased specificity for the gene of interest.

### 2.5. Measurement of absolute copy number of mRNAs in RT-qPCR assays

We developed preamplification enhanced real time quantitative polymerase chain reaction (RT-qPCR) assay to compensate for low RNA yield from urinary cells (Muthukumar et al., 2005; Abuhelaiqa et al., 2021). In the preamplification step, the cDNA was pre-amplified with gene specific primer pairs in a final reaction volume of 10.0 μl in a 0.2 ml PCR tube and each sample contained 3.0 μl cDNA, 5.0μl Platinum^®^ Multiplex PCR Master Mix, 1.68 μl primer mix (50 μM sense and 50 μM antisense primer per gene) and 0.32 μl water to a final volume of 10 μl. We used Platinum Multiplex PCR Master mix in the current study for multiplexing 20 amplicons and the flexibility it offered in terms of PCR product size. Following vortexing, the PCR amplification was set up in a Veriti thermal cycler (Applied Biosystems) and the PCR profile consisted of an initial hold at 95 °C for 2 min, 11 cycles of denaturing at 95 °C for 30 seconds, primer annealing at 60 °C for 90 seconds and extension at 72 °C for 1 min., and final extension at 72 °C for 10 min and final hold at 4 °C. The PCR profile used in the current study enabled amplification of 20 different mRNAs with a near perfect amplification efficiency. At the end of 11 cycles of amplification, we diluted the PCR amplicons by adding 290 μl of TE buffer to the 10 μl PCR reaction. For quantification of mRNAs, the RT-qPCR assay was performed on the Applied Biosystems QuantStudio™ 6 Real-Time PCR system (ThermoFisher) using 2.5 μl of diluted PCR amplicons in a final reaction volume of 20 μl. In the CTOT-04 study, mRNAs were quantified using the Applied Biosystems 7500 Fast or 7900 HT Real Time PCR system (Suthanthiran et al., 2013). A platform comparison study by the PCR system manufacturer found equivalency among these three instruments with a 2-fold difference being detected with a 99.9% confidence with any one of the three PCR systems.

We calculated absolute copy numbers of mRNAs using our previously described Bak standard curve for absolute quantification in RT-qPCR assays (Suthanthiran et al., 2013). We established the Bak standard curve with 6 log_10_ concentrations of Bak amplicon (starting copy number 2.5 × 10^6^) as the template and PCR amplification with Bak specific primer pair and detection using Bak specific TaqMan probe. The amplification efficiencies of Bak and CD3ε, CXCL10, TGFβ1 and 18S rRNA were greater than 90% in the RT-qPCR assays. Thus, there was minimal bias due to different amplification efficiencies of the target genes and the Bak amplicon.

### 2.6. Data analysis

Mean, median and the 25^th^ percentile and the 75^th^ percentile values were calculated for continuous variables. Non-normally distributed data were analyzed using non-parametric tests. Absolute copy number for each mRNA was analyzed before and after normalization with 18S rRNA copies (×10^-6^) and with and without log_10_-transformation. Area under the receiver operating characteristic curve (AUROC) was used to analyze the diagnostic (discriminating) ability of mRNA copy numbers. The CTOT-04 three gene diagnostic score was calculated using the previously reported logistic regression equation: -6.1487+0.8534 log_10_ (CD3ε/18S)+0.6376 log_10_ (CXCL10/18S)+1.6464 log_10_ (18S) (Suthanthiran et al., 2013). In this equation, -6.1487 is the intercept, and 0.8534, 0.6376, and 1.6464 are the slopes (coefficients), respectively, for the log_10_ (CD3ε/18S), log_10_ (CXCL10/18S), and log_10_ (18S) values in the best-fitting logistic-regression model. The units of measurement in the RT-qPCR assays for CD3ε mRNA and CXCL10 mRNA are copy number per microgram of total RNA, and the units for 18S rRNA are number of copies (×10^-6^) per microgram of total RNA. Our previously validated TCMR diagnostic cut point of -1.213 for the CTOT-04 three gene diagnostic score (Suthanthiran et al., 2013) was used to calculate the sensitivity and specificity for predicting TCMR in the current study. Our previously validated cut point of 6.5×10^8^ BKVN VP1 mRNA copy number per microgram of total RNA (Dadhania et al., 2010) was used to calculate sensitivity and specificity for predicting BKVN in the current study.

We used GraphPad Prism 9 (Version 9.4.1 for Mac) statistical software package for data analysis.

## 3. Results

### 3.1. Kidney allograft study cohorts and biopsy findings

Table 1 is a summary of the characteristics of the kidney allograft recipients profiled to determine the diagnostic performance characteristics of WCHP. Donor information is also included in Table 1.

**Table 1.** Characteristics of kidney allograft recipient study cohorts.

| Characteristics | No Rejection Cohort | TCMR Cohort | BKVN Cohort |
| --- | --- | --- | --- |
| Number of kidney allograft recipients <sup>a</sup> | 29 | 11 | 7 |
| Number of urine samples <sup>a</sup> | 29 | 12 | 7 |
| *At the time of kidney transplantation* |  |  |  |
| **Donor information** |  |  |  |
| Age (years), mean (SD) | 45.2 (14.3) | 46.5 (11.5) | 50 (7.2) |
| Female, n (%) | 16 (57.1) | 8 (66.7) | 4 (57.1) |
| Race |  |  |  |
| Black, n (%) | 4 (15.4) | 1 (8.3) | 1 (14.2) |
| Other, n (%) | 5 (19.2) | 8 (66.7) | 3 (42.9) |
| White, n (%) | 17 (65.4) | 3 (25.0) | 3 (42.9) |
| Deceased donor, n (%) | 10 (34.5) | 4 (33.3) | 5 (71.4) |
| **Recipient information** |  |  |  |
| Age (years), mean (SD) | 48.8 (12.9) | 49 (16.3) | 52 (11.0) |
| Female, n (%) | 12 (41.4) | 5 (41.7) | 4 (57.1) |
| Race |  |  |  |
| Black, n (%) | 7 (24.1) | 4 (33.3) | 3 (42.9) |
| Other, n (%) | 7 (24.1) | 5 (41.7) | 2 (28.6) |
| White, n (%) | 15 (51.7) | 3 (25.0) | 2 (28.6) |
| Pretransplant DSA <sup>b</sup> |  |  |  |
| Data available, n (%) | 26 (89.7) | 11 (91.7) | 7 (100) |
| DSA positive, n (%)<sup>c</sup> | 11 (42.3) | 2 (18.2) | 3 (42.9) |
| DSA against HLA Class I, n (%) | 9 (34.6) | 1 (9.1) | 3 (42.9) |
| DSA against HLA Class II, n (%) | 6 (23.1) | 1 (89.1) | 0 (0) |
| Induction therapy |  |  |  |
| Antithymocyte globulin (ATG), n (%) | 17 (63.0) | 9 (75.0) | 6 (85.7) |
| Basiliximab, n (%) | 4 (14.8) | 1 (8.3) | 1 (14.2) |
| ATG+ Basiliximab, n (%) | 6 (22.2) | 2 (16.7) | 0 (0) |
| *At the time of index kidney allograft biopsy* |  |  |  |
| For-cause (clinically indicated) biopsy, n (%) | 18 (62.1) | 12 (100) | 7 (100) |
| Time from transplantation to kidney allograft biopsy, days, median (IQR) | 387 (1642) | 286 (971) | 126 (111) |
| Serum creatinine, mg/dL, median (IQR) | 1.72 (1.5) | 3.06 (1.6) | 2.00 (1.3) |
| DSA associated with index allograft biopsy |  |  |  |
| Data available, n (%) | 22 (75.9) | 11 (91.7) | 7 (100) |
| DSA positive, n (%) | 11 (50) | 4 (36.4) | 1 (14.2) |
| DSA against HLA Class I, n | 9 (40.9) | 3 (27.3) | 1 (14.2) |
| DSA against HLA Class II, n | 6 (27.3) | 3 (27.3) | 0 (0) |
| Total RNA isolated from urine using WCHP |  |  |  |
| Yield, median (µg) | 0.81 | 2.07 | 1.83 |
| Purity, median (A260/280) | 1.95 | 1.98 | 1.97 |

<sup>a</sup>The Table shows a total of 48 urine samples from 45 unique kidney transplant recipients since one patient with TCMR biopsy diagnosis provided 2 urine samples, and 2 urine samples were from two patients with biopsies showing TCMR and IHC positive for SV40/BKV large T antigen are included under both TCMR and BKVN cohorts.
<sup>b</sup>Donor HLA Specific IgG Antibodies (DSA) detected in the kidney allograft recipient using LABScreen single antigen (LSA) class I assay (LS1A04) and the LSA class II assay (LS2A01), respectively (One Lambda).
<sup>c</sup>DSA with mean fluorescence intensity (MFI) > 1000 was considered positive.
<sup>d</sup>Total RNA from biopsy matched urine samples were isolated using the WCHP and yield (absorbance at A260) and purity (ratio of A260 to A280) were measured using Nanodrop One Spectrophotometer.

Photomicrographs of kidney allograft biopsies stained with hematoxylin and eosin (left panel), with PAS (middle panel) and stained for BKV large T antigen using the PAb416 mouse mAb (right panel) are shown in Fig. 1. A representative No Rejection (NR) biopsy negative for histologic hallmarks of TCMR, AMR and negative for intragraft SV40/BKV large T antigen is shown in Fig. 1a. A representative TCMR biopsy with tubulitis (t) and interstitial infiltration (i) but negative for BKV large T antigen is shown in Fig. 1b. An illustrative kidney allograft biopsy positive for intragraft SV40/BKV large T antigen, tubular cytopathy and also for graft inflammation is shown in Fig. 1c.

**Fig. 1.**
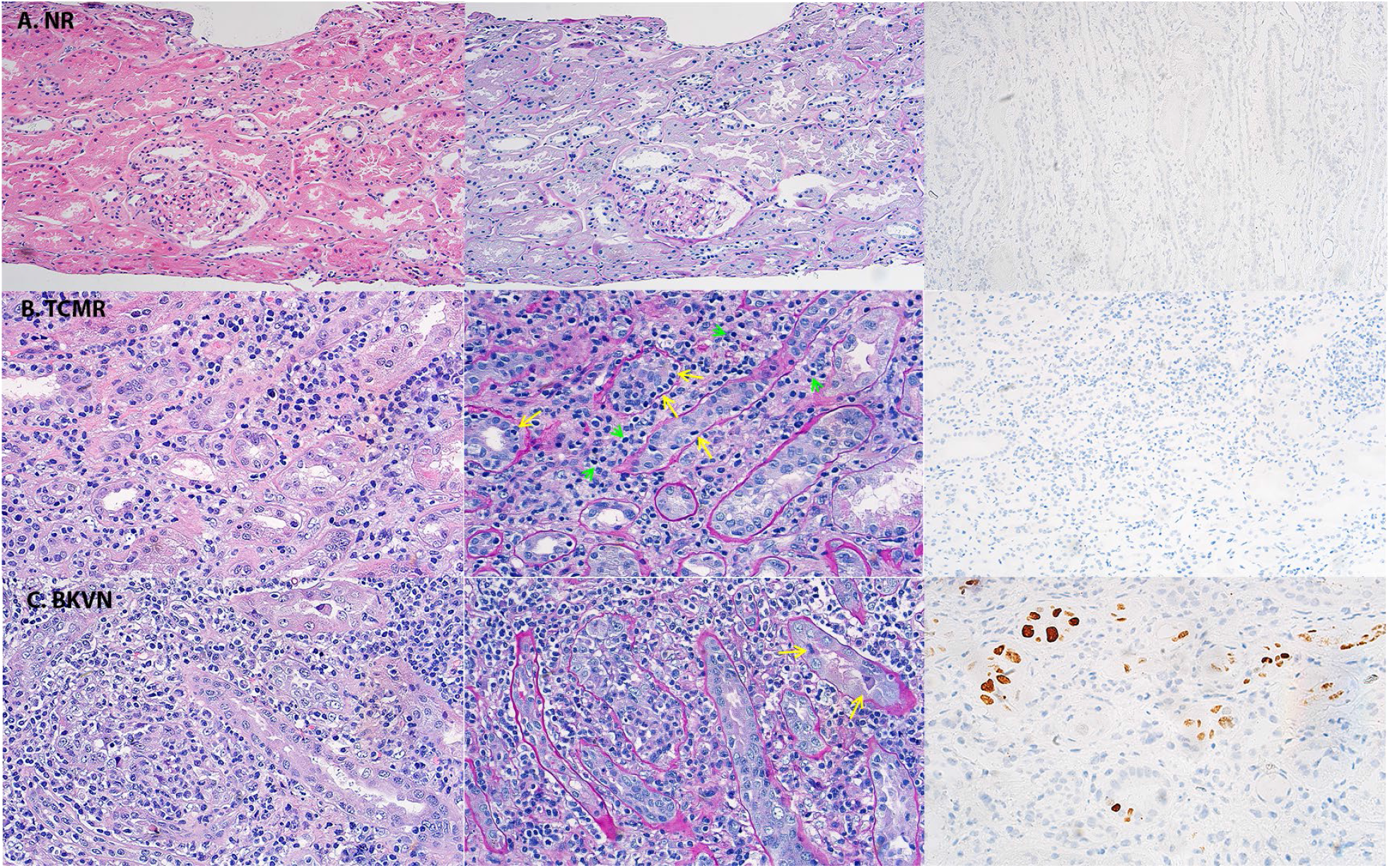
Representative kidney allograft biopsies classified as No Rejection biopsy, TCMR biopsy or BKVN biopsy. Photomicrographs of kidney allograft biopsies stained with hematoxylin and eosin (left panel), with PAS (middle panel) (magnification 400x) and immunohistochemistry (IHC) for SV40/BKV large T antigen using the PAb416 mouse mAb (right panel). A. A representative No Rejection biopsy (NR) negative for histologic hallmarks of TCMR, AMR and IHC negative for SV40/ BKV large T antigen. B. A representative TCMR biopsy with tubulitis (yellow arrows) and interstitial infiltration (green arrows) but negative for SV40/BKV large T antigen. C. An illustrative BKVN kidney allograft biopsy with viral cytopathic changes in the tubules (yellow arrows) and IHC positive for intranuclear SV40/BKV large T antigen. The instructive BKVN biopsy also shows the concurrent presence of intragraft inflammation often present in biopsies diagnosed as BKVN.

The Banff kidney allograft biopsy classification schema applies a semiquantitative scoring system of 0, 1, 2 or 3 for scoring acute and chronic histological lesions within the kidney allograft. Fig. 2 is a heatmap of the histological lesions using the Banff semiquantitative scale for kidney allograft biopsy findings. All No Rejection biopsies (N1 to N29) did not meet the Banff criteria for TCMR or AMR, but two biopsies (N18 and N24) met the criteria for Borderline changes based on Banff scores of t1 and i1. All 29 No Rejection biopsies were negative for intragraft SV40/BKV large T antigen and 28 of 29 biopsies were negative for intragraft deposition of complement 4 split product d (C4d neg), and the remaining one biopsy (N14) stained weakly positive (<25%) for C4d. The median and interquartile range (IQR) time from transplantation to No Rejection biopsy was 387 (1642) days, and, as expected, several of the No Rejection biopsies showed chronic vascular (cv) changes, tubular atrophy (ct), and/or interstitial fibrosis (ci).

**Fig. 2.**
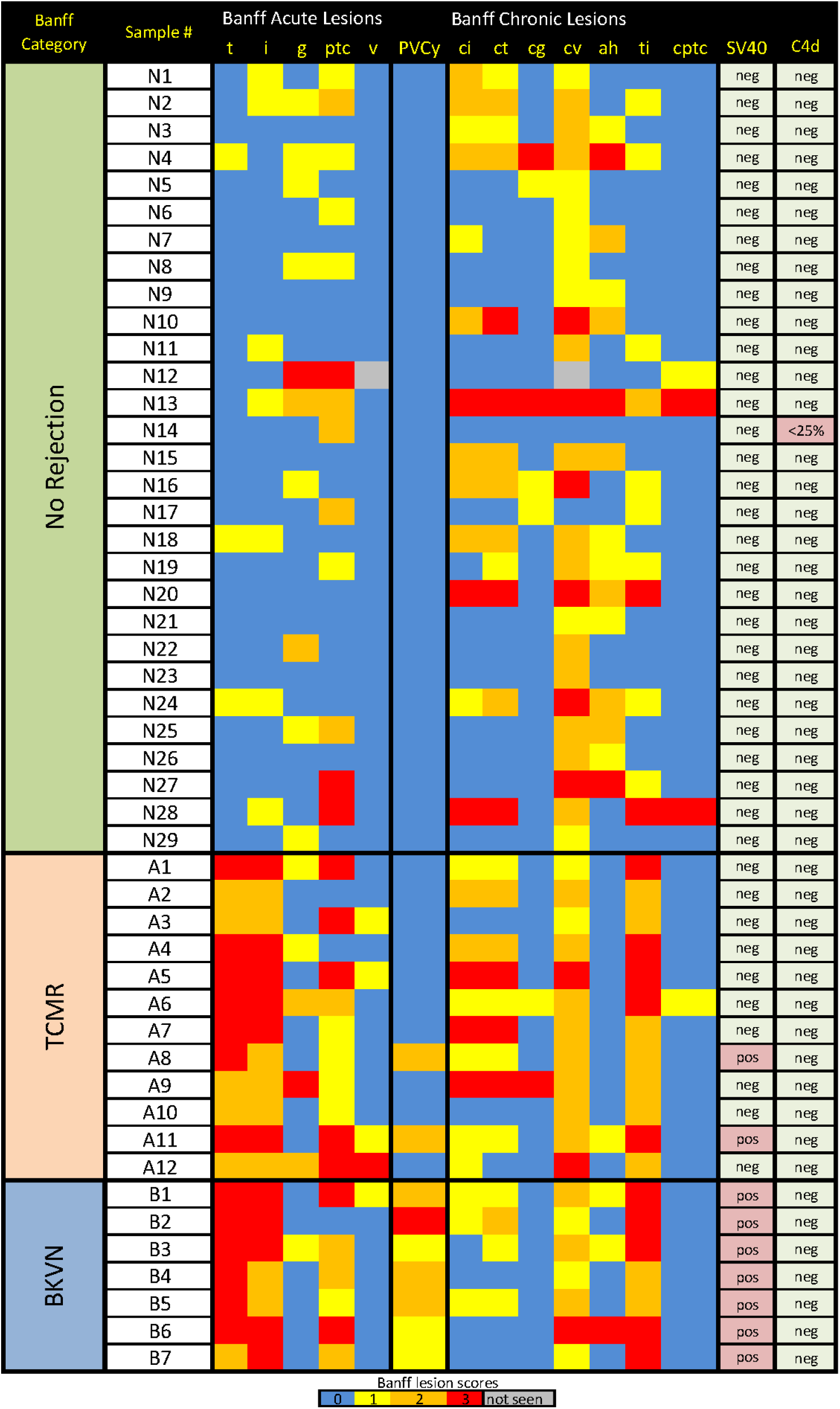
Heatmap of the histological lesions in kidney allograft biopsies scored using the Banff semi-quantitative scale for kidney allograft biopsy findings. Banff acute scores are shown for tubulitis (t), interstitial inflammation (i), glomerulitis (g), peritubular capillary inflammation (ptc), and vascular inflammation (v). Banff chronic scores for interstitial fibrosis (ci), tubular atrophy (ct), chronic glomerulopathy (cg), chronic vascular lesions (cv), arteriolar hyaline thickening (ah), total interstitial inflammation (ti) and peritubular capillary inflammation (cptc) are also shown for each biopsy. IHC for SV40/BKV large T antigen (SV40) and intragraft deposition of complement factor 4d (C4d) are also included in the heatmap along with polyoma virus cytopathic score (PVCy) (Nickeleit et al., 2020). All No Rejection biopsies (N1 to N29) did not meet the Banff criteria for TCMR or AMR, but two biopsies (N18 and N24) met the criteria for Borderline changes based on Banff scores of t1 and i1. All 29 No Rejection biopsies were negative (neg) for intragraft SV40/BKV large T antigen and 28 of 29 biopsies were also negative for C4d and one biopsy (N14) had minimal (<25%) C4d deposit. The No Rejection biopsies also showed concurrent chronic changes. All TCMR biopsies (A1 to A12) met Banff criteria for TCMR with tubulitis score of 2 or greater and interstitial infiltration score of 2 or greater, and were all negative C4d, a histologic hallmark of AMR. The TCMR biopsies also showed concurrent chronic changes. Two of the TCMR biopsies (A8 and A11) were positive (pos) for SV40/BKV large T antigen and these biopsies are also included in the BKVN group. All biopsies classified as BKVN (B1 to B7) were positive for intragraft SV40/BKV large T antigen and negative for C4d. The heatmap shows polyomavirus cytopathic (PVCy) score for each of the biopsies; a score of 0 (blue) is assigned when the biopsy shows no viral inclusion bodies and negative for IHC for SV40/BKV large T antigen.

All TCMR biopsies (A1 to A12) met Banff criteria for TCMR with tubulitis score of 2 or greater and interstitial infiltration score of 2 or greater, and a histologic hallmark of AMR, intragraft C4d, was negative in each instance. The median and IQR time from transplantation to TCMR biopsy was 286 (971) days and, as expected, displayed cv, ct, and/or ci. As shown, 2 of the 12 TCMR biopsies (A8 and A11) were positive for SV40/BKV large T antigen, and these 2 biopsies are included in the TCMR group and in the BKVN group.

All biopsies classified as BKVN biopsies (B1 to B7) were positive for intragraft SV40/BKV large T antigen and all were negative for C4d. BKVN is often associated with intragraft inflammation and chronic changes, and this is reflected in the high acute and chronic scores in the BKVN biopsies. The concurrence of intragraft inflammation with BKVN is reflected in BKVN classification schema with BKVN A representing BKVN without any inflammation, BKVN B with inflammation and BKVN C with fibrosis (Drachenberg et al., 2004). The Heatmap also shows polyoma virus associated cytopathic (PVCy) (Nickeleit et al., 2020). A score of 0 was observed in all biopsies that were negative for SV40/BKV large T antigen staining by immunohistochemistry.

### 3.2. Noninvasive diagnosis of TCMR using the WCHP

We used the WCHP to isolate total RNA from the biopsy matched urine samples from the kidney allograft recipients and the yield and the high purity of the total RNA isolated with this protocol are shown in Table 1. We measured absolute copy numbers of mRNAs in the total RNA isolated from biopsy matched urine samples from the kidney allograft recipients. Violin plots portraying the distribution of absolute copy numbers of mRNAs in the urine samples matched to TCMR biopsies or No Rejection biopsies are shown in Fig. 3. In accord with the CTOT-04 study findings, absolute copy numbers of CD3ε mRNA, CXCL10 mRNA, TGFβ1 mRNA and 18S rRNA were higher in the TCMR group than the No Rejection group. The median (25^th^ percentile and 75^th^ percentile) CD3ε mRNA copy number was 3530 (487, 6178) per microgram of total RNA in the 12 urines matched to 12 TCMR biopsies and 191 copies (31, 743) per microgram of total RNA in the 29 urines matched to the 29 No Rejection biopsies (Fig. 3A, P=0.0004, Mann Whitney test). CXCL10 mRNA copy number was 3395 (322, 5460) copies in the TCMR group and 151 (70, 973) in the No Rejection biopsy group (Fig. 3B, P=0.001). TGFβ1 mRNA copy number was 13200 (7855, 35025) copies in the TCMR biopsy group and 4710 (476, 16900) in the No Rejection biopsy group (Fig. 3C, P=0.02). 18S rRNA copy number was 1.73×10^9^ (1.15×10^9^, 8.57×10^9^) in the TCMR biopsy group and 8.01×10^8^ (2.97×10^8^, 3.32×10^9^) in the No Rejection biopsy group (Fig. 3D, P=0.04)

**Fig. 3.**
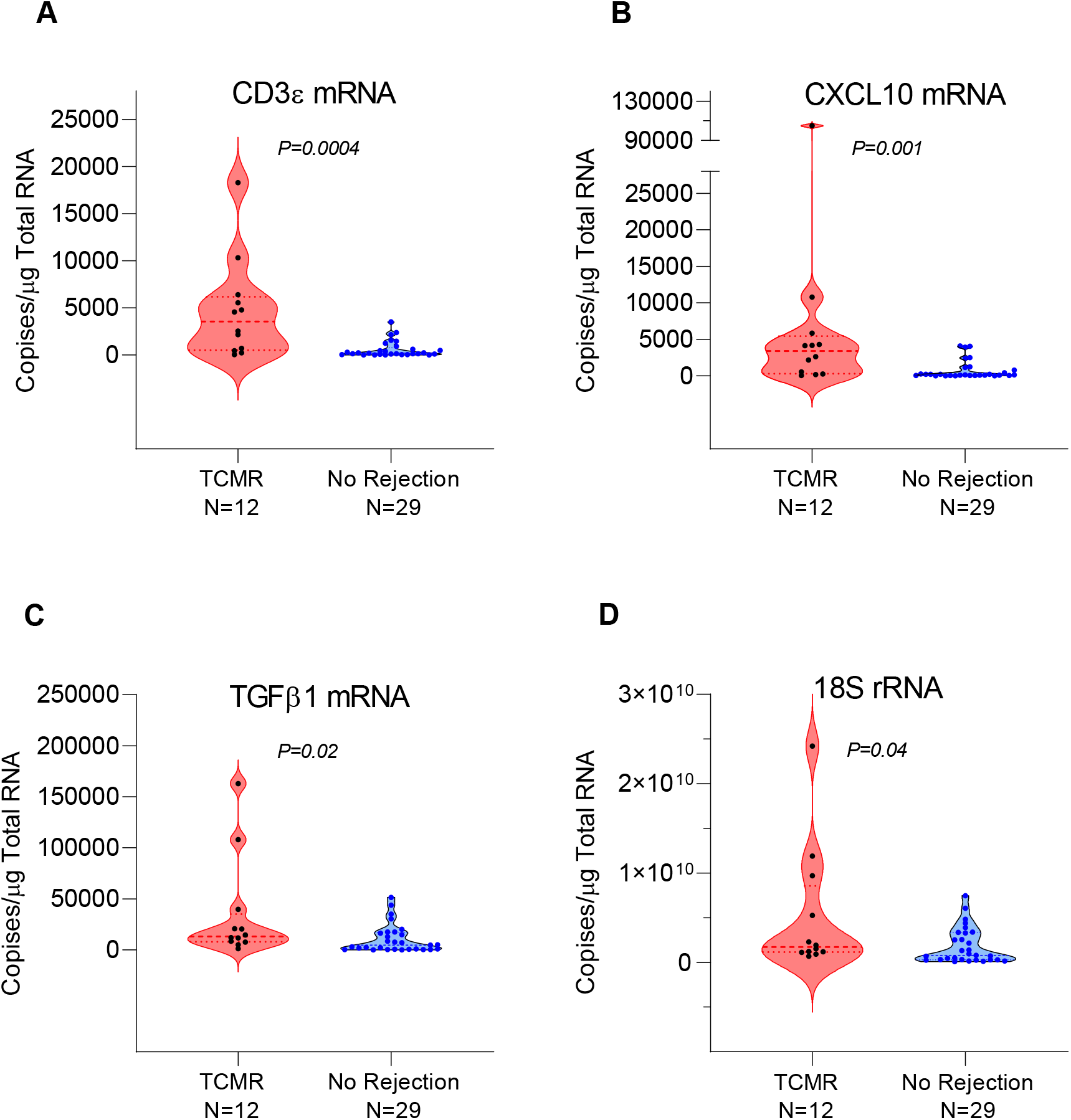
Absolute copy numbers of mRNAs per microgram of total RNA isolated from biopsy matched urine samples from kidney allograft recipients. Total RNA was isolated from urine samples using the WCHP and absolute copy numbers of mRNAs were quantified using the RT-qPCR assays. Violin plots show minimum, 25^th^, 50^th^ (median), 75^th^ quartiles for CD3ε mRNA (A), CXCL10 mRNA (B), TGFβ1 mRNA (C), and 18S rRNA (D) in biopsy matched urine samples from unique kidney allograft recipients. The levels of CD3ε, CXCL10, TGFβ1 mRNAs, and 18S rRNA were significantly higher in urine matched to TCMR biopsies (n=12 biopsy matched urine samples from 11 patients) compared to urine matched to No Rejection (NR) biopsies (N=29 biopsy matched urine samples from 29 patients). P values calculated using Mann-Whitney test.

The CTOT-04 three gene diagnostic signature score was higher in the TCMR biopsy group than in the No Rejection biopsy group; the median score was -0.448 (-1.664, 0.204) in the TCMR biopsy group and -2.542 (-3.267, -1.365) in the No Rejection biopsy group (Fig. 4A, P=0.0005).

**Fig. 4.**
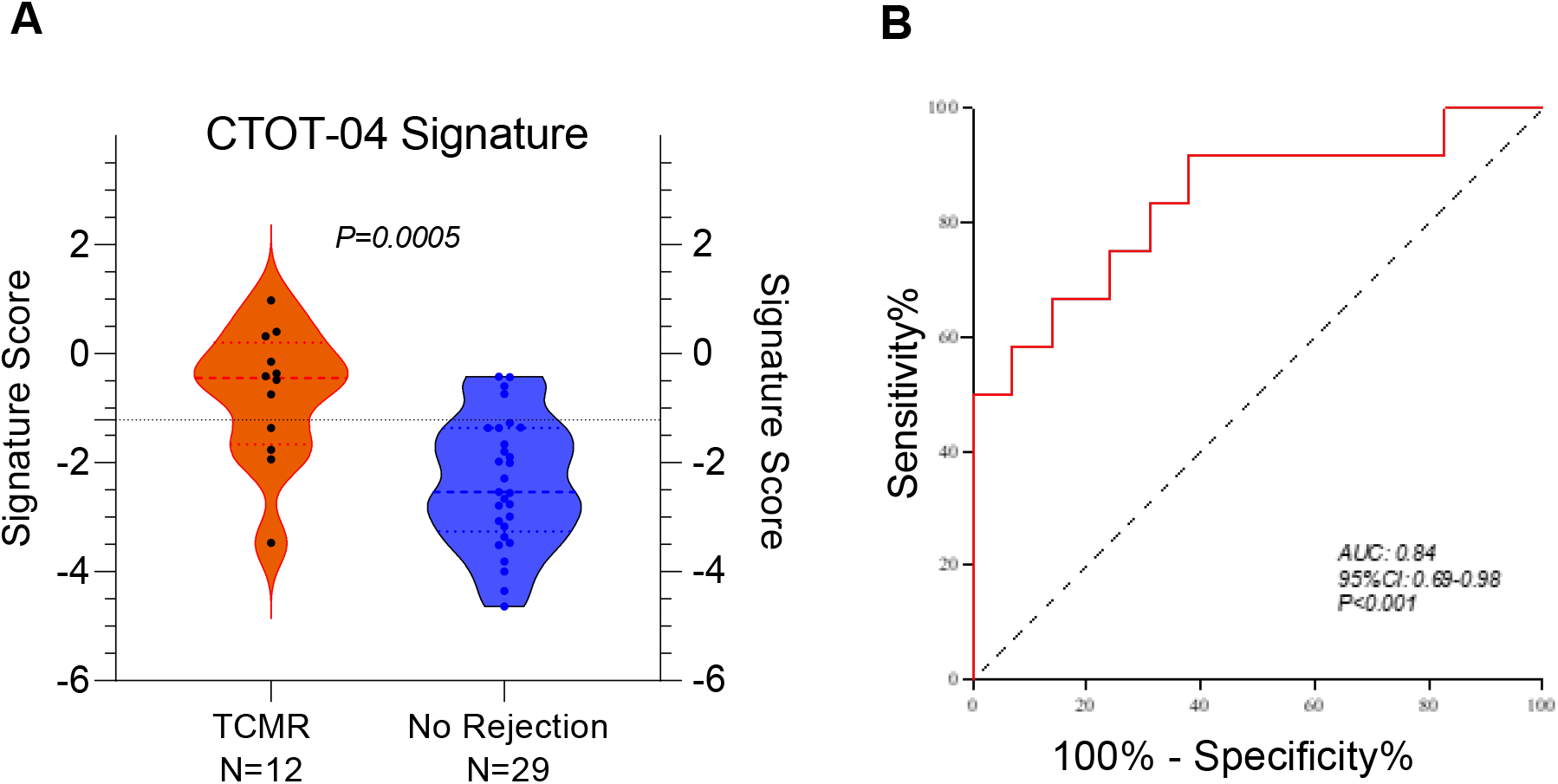
CTOT-04 three gene TCMR diagnostic signature score in total RNA isolated using the WCHP is diagnostic of TCMR. A. Violin plots of the CTOT-04 three gene diagnostic signature score (calculated using logistic regression equation in Suthanthiran et al. 2013) in the total RNA isolated from biopsy matched urines using WCHP. The median (25^th^ percentile, 75^th^ percentile) score was -0.448 (-1.664, 0.204) in the TCMR biopsy group (n=12 biopsy matched urine samples from 11 patients) and -2.542 (-3.267, -1.365) in the No Rejection biopsy group (N=29 biopsy matched urine samples from 29 patients). (P=0.0005, Mann Whitney test). The line across the violin plots indicates the -1.213 diagnostic cutpoint from the CTOT-04 study (Suthanthiran et al., 2013). B. ROC curve analysis of the CTOT-04 three gene TCMR diagnostic signature score in urines processed using the WCHP discriminated patients with TCMR biopsies from those with No Rejection biopsies with an AUROC of 0.84 (95% Confidence Intervals [CI], 0.69 to 0.98) (P<0.001). An AUROC 1.0 is a perfect discriminator and 0.5 is no better than chance.

ROC curve analysis of the CTOT-04 three gene diagnostic signature score in urines processed using the WCHP discriminated patients with TCMR biopsies from those with No Rejection biopsies with an AUROC of 0. 84 (95% Confidence Intervals [CI], 0.69 to 0.98) (P<0.001) (Fig. 4B). With the previously identified CTOT-04 cutpoint of -1.213 for optimizing sensitivity and specificity (Suthanthiran et al., 2013), the sensitivity for diagnosing TCMR in the current study was 67% (95% CI, 35 to 89) and the specificity was 86% (95% CI, 67 to 95) (Table 2, P=0.0016, Fischer exact test).

**Table 2.** CTOT-04 Three-Gene Score in total RNA isolated using WCHP is diagnostic of TCMR.

| **CTOT-04 TCMR Diagnostic Signature Score Cutpoint<sup>a</sup>** | **Kidney Allograft Biopsy<sup>b</sup>** |  |
| --- | --- | --- |
|  | **TCMR<sup>c</sup>**<br>**(N=12)** | **No Rejection<sup>c</sup>**<br>**(N=29)** |
| > -1.213 | 8 | 4 |
| ≤ -1.213 | 4 | 25 |
| Fisher exact test |  | P=0.0016 |
| Sensitivity (95% confidence interval) | 67% (35% to 89%)<sup>a</sup> |  |
| Specificity (95% confidence interval) |  | 86% (67% to 95%)<sup>a</sup> |

<sup>a</sup> CTOT-04 urinary cell three gene signature score -1.213 optimized sensitivity and specificity for diagnosing TCMR in the CTOT-04 study (Suthanthiran et al., 2013). In the current study, the logistic regression equation used in the CTOT-04 study was used to calculate the three gene TCMR diagnostic signature score in the 41 urine samples matched to 41 biopsies from 40 patients and the -1.213 cutpoint that maximized sensitivity and specificity for diagnosing TCMR in the CTOT-04 study was used to classify urine samples and calculate sensitivity and specificity for predicting TCMR in the current study.
<sup>b</sup> 40 kidney allograft recipients underwent 41 percutaneous needle core biopsies, and the 41 biopsies were classified using the Banff classification schema (Loupy et al., 2017).
<sup>c</sup> Each biopsy classified as TCMR met Banff category TCMR Grade 1 or greater. Two of the 12 TCMR biopsies were also positive for intra-graft SV40/BKV large T antigen. The No Rejection
biopsies did not display histological features of TCMR or AMR and were all IHC negative for intragraft SV40/BKV large T antigen.

Statistical analysis of the AUROC (P>0.05, Wilcoxon test), sensitivity (P=0.44, Fisher’s exact test to compare proportions) and specificity (P=0.33, Fisher’s exact test to compare proportions) observed in the current WCHP-based current study and the earlier CTOT-04 study showed no significant differences in any of the assessed diagnostic parameters between the two studies.

### 3.4. Noninvasive diagnosis of BKVN using the WCHP

Demographics of the kidney allograft recipients diagnosed with BKVN biopsies are also summarized in Table 1. We measured, using RT-qPCR assays, the absolute copy number of urinary cell BKV VP1 mRNA in urines matched to kidney allograft biopsies diagnosed as BKVN and in urines matched to kidney allograft biopsies diagnosed as No Rejection biopsies and negative for intragraft SV40/BKV large T antigen. We also measured absolute copy number of 18S rRNA copy number in these biopsy matched urine samples.

As expected from our earlier studies (Ding et al., 2002; Dadhania et al., 2010), urinary cell BKV VP1 mRNA copy number in the urines matched to kidney allograft biopsies diagnosed as BKVN was significantly higher than in the urines matched to No Rejection biopsies. The median (25^th^ percentile, 75^th^ percentile) BKV VP1 mRNA copy number was 1.67 ×10^10^ (1.75 ×10^9^, 4.81 ×10^10^) per microgram of total RNA in the 7 urines matched to 7 BKVN biopsies and 0 (0, 585) per microgram of total RNA in the urines 29 matched to the 29 No Rejection biopsies (Fig. 5a, P<0.0001, Mann Whitney test). Activated cells express higher level of 18S rRNA compared to resting cells (Roge et al., 2007). Urinary cell level of 18S rRNA was higher in the BKVN group than in the No Rejection group; the median (25^th^ percentile, 75^th^ percentile) 18S rRNA copy number was 6.90×10^9^ (3.75 ×10^9^, 9.47 ×10^9^) per microgram of total RNA in 7 urines matched to the 7 BKVN biopsies and 8.01 ×10^8^ in the 29 urines matched to the 29 No Rejection biopsies (P=0.001, Mann Whitney test).

**Fig. 5.**
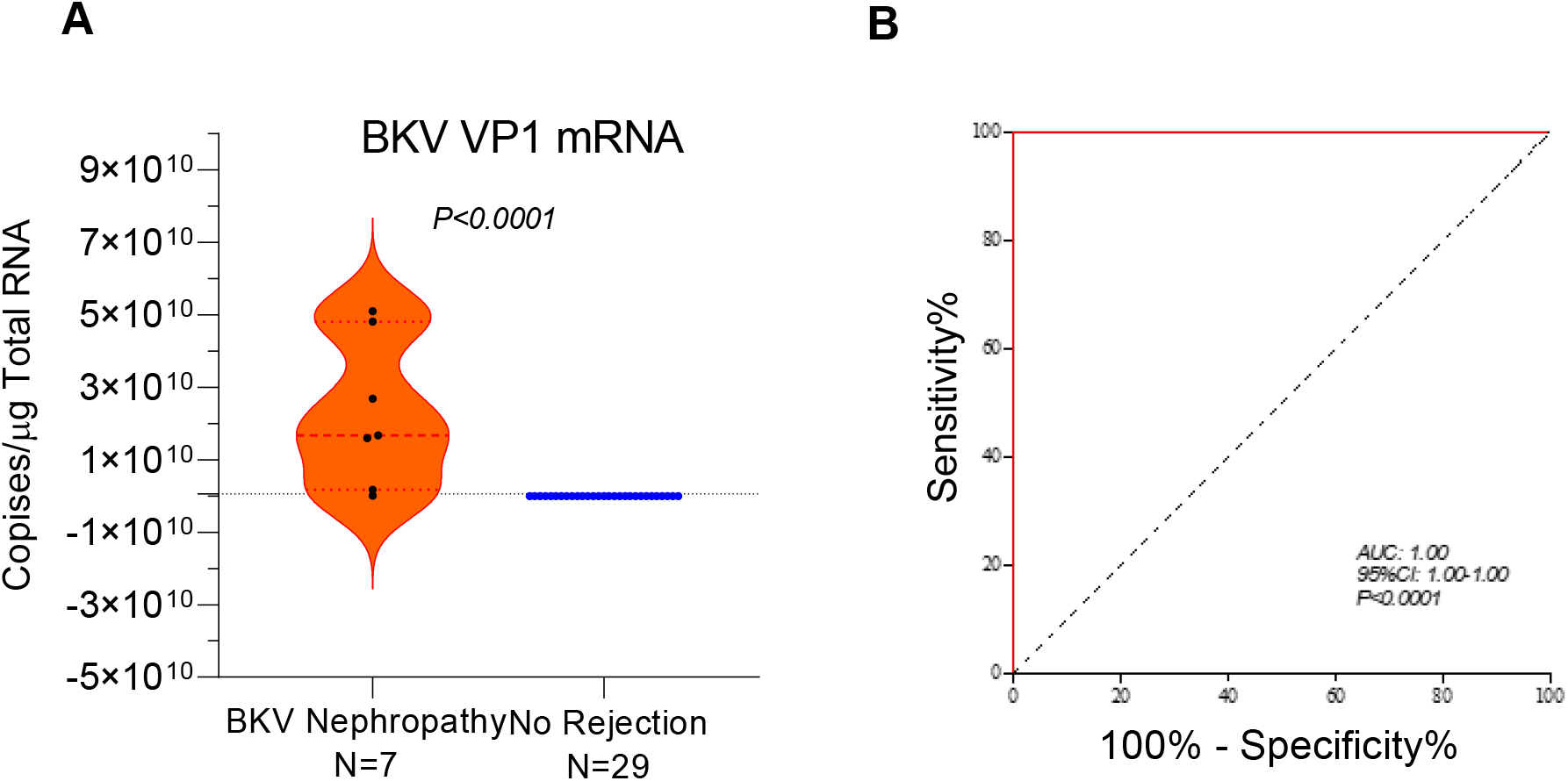
BKV VP1 mRNA copy number in total RNA isolated using WCHP is diagnostic of BKVN. A. Violin plots of BKV VP1 mRNA copy numbers in total RNA isolated using WCHP. The median (25^th^ percentile, 75^th^ percentile) BKV VP1 mRNA copy number was 1.67×10^10^ (1.75×10^9^, 4.81×10^10^) per microgram of total RNA in the 7 urines matched to 7 BKVN biopsies and 0 (0, 585) per microgram of total RNA in the 29 urines matched to the 29 No Rejection biopsies (P<0.0001, Mann Whitney test). B. ROC curve analysis of the BKV VP1 mRNA copy numbers in urines processed using the WCHP perfectly discriminated patients with BKVN biopsies from those with No Rejection with an AUROC of 1.000 (95% CI, 1.000 to 1.000, P<0.0001).

ROC curve analysis of the BKV VP1 mRNA copy numbers in urines processed using the WCHP perfectly discriminated patients with BKVN biopsies from those with No Rejection with an AUROC of 1.000 (95% CI, 1.000 to 1.000, P<0.0001) (Fig. 5b). In our earlier urinary cell BKV VP1 mRNA copy number validation study, the Youden cutpoint value that maximized sensitivity and specificity for the noninvasive diagnosis of BKVN was 6.5 × 10^8^ BKV VP1 mRNA copies per microgram of total RNA isolated from urinary cells (Dadhania et al., 2010). In the current study, the sensitivity was 86% (95% CI, 42 to 99) and the specificity was 100% (95% CI, 85 to 100) with the validated cutpoint of 6.5 × 10^8^ BKV VP1 mRNA copies per microgram of total RNA (Table 3, P<0.0001, Fischer exact test).

**Table 3.**
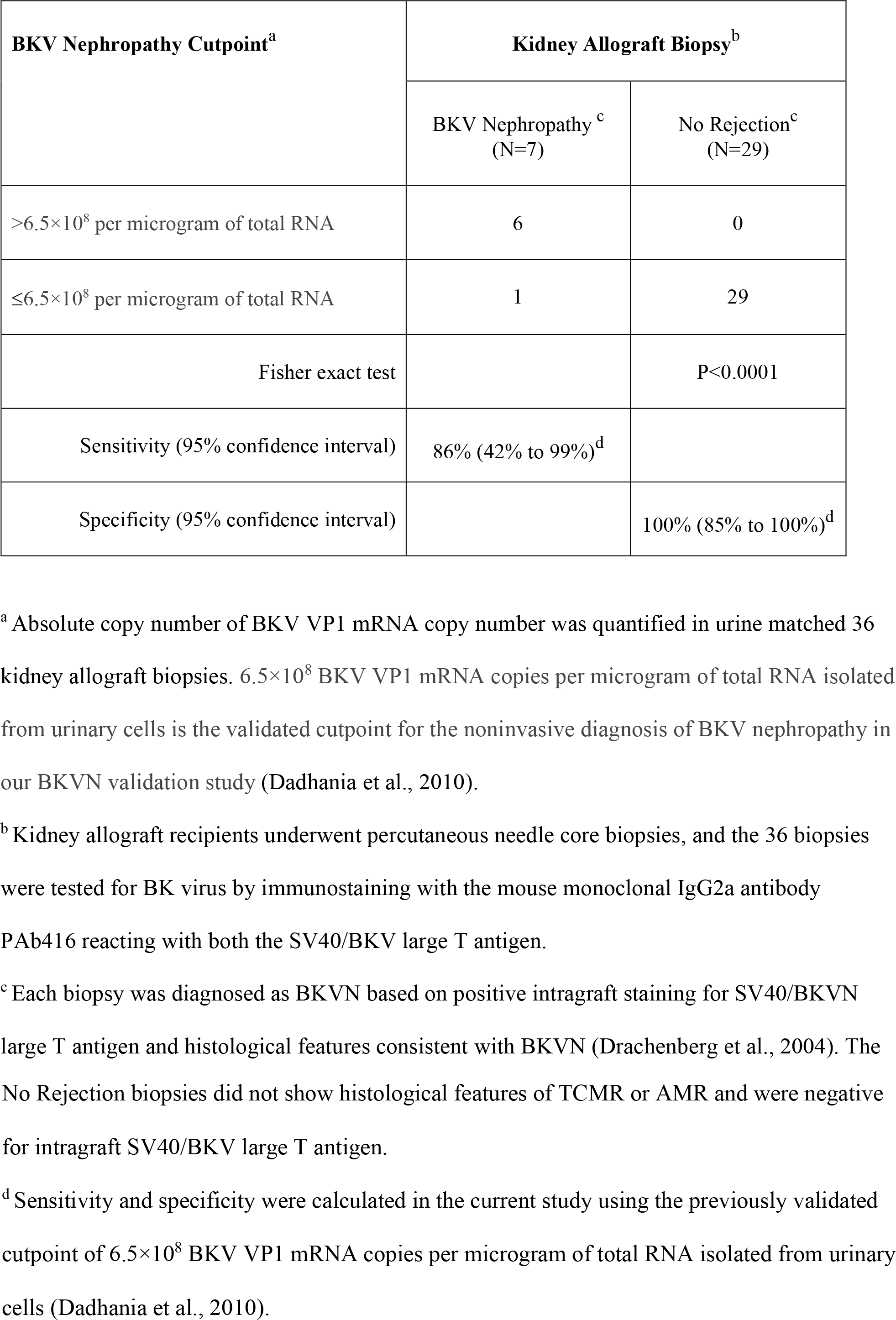
Noninvasive diagnosis of BKVN by measurement of urinary cell BKV VP1 mRNA copy number in total RNA isolated using WCHP.

| **BKV Nephropathy Cutpoint<sup>a</sup>** | **Kidney Allograft Biopsy<sup>b</sup>** |  |
| --- | --- | --- |
|  | **BKV Nephropathy <sup>c</sup>**<br>**(N=7)** | **No Rejection<sup>c</sup>**<br>**(N=29)** |
| >6.5×10<sup>8</sup> per microgram of total RNA | 6 | 0 |
| ≤6.5×10<sup>8</sup> per microgram of total RNA | 1 | 29 |
| Fisher exact test |  | P<0.0001 |
| Sensitivity (95% confidence interval) | 86% (42% to 99%)<sup>d</sup> |  |
| Specificity (95% confidence interval) |  | 100% (85% to 100%)<sup>d</sup> |

<sup>a</sup> Absolute copy number of BKV VP1 mRNA copy number was quantified in urine matched 36 kidney allograft biopsies. 6.5×10<sup>8</sup> BKV VP1 mRNA copies per microgram of total RNA isolated from urinary cells is the validated cutpoint for the noninvasive diagnosis of BKV nephropathy in our BKVN validation study (Dadhania et al., 2010).
<sup>b</sup> Kidney allograft recipients underwent percutaneous needle core biopsies, and the 36 biopsies were tested for BK virus by immunostaining with the mouse monoclonal IgG2a antibody PAb416 reacting with both the SV40/BKV large T antigen.
<sup>c</sup> Each biopsy was diagnosed as BKVN based on positive intragraft staining for SV40/BKVN large T antigen and histological features consistent with BKVN (Drachenberg et al., 2004). The
No Rejection biopsies did not show histological features of TCMR or AMR and were negative for intragraft SV40/BKV large T antigen.
<sup>d</sup> Sensitivity and specificity were calculated in the current study using the previously validated cutpoint of $6.5 \times 10^8$ BKV VP1 mRNA copies per microgram of total RNA isolated from urinary cells (Dadhania et al., 2010).

Statistical analysis of the AUROC (P>0.05, Wilcoxon test), sensitivity (P=0.33, Fisher’s exact test to compare proportions) and specificity (P=0.36, Fisher’s exact test to compare proportions) observed in the current WCHP based current investigation and our earlier BKVN validation study showed no significant differences in any of the assessed diagnostic parameters between the two studies.

## 4. DISCUSSION

We recently developed the ZFBP to eliminate centrifugation of the urine sample to prepare urinary cell pellet as well as to obviate the need for an RNA preservative, refrigeration at -80°C and shipment in cold containers to a gene expression profiling laboratory for urinary cell mRNA profiling (Snopkowski et al., 2021). A distinct advantage of the ZFBP is that the kidney allograft recipients could be trained to perform the initial urine processing steps at home and post the urinary cell lysate containing RNA to a gene expression profiling laboratory at ambient temperature. In the current investigation, we retained the advantages of the ZFBP but modified the procedure to enrich for mRNAs in the total RNA isolated from the urine sample. Results summarized in this report demonstrate that noninvasive diagnosis of TCMR with performance characteristics observed in the CTOT-04 study is feasible using the WCHP, and noninvasive diagnosis of BKVN with performance characteristics observed in the BKVN validation study is feasible using the WCHP.

In the CTOT-04 study, the parsimonious three gene diagnostic signature of 18S rRNA normalized CD3ε mRNA, 18s rRNA normalized CXCL10 mRNA, and 18S rRNA discriminated kidney allograft recipients with TCMR biopsies from those without TCMR biopsies and the AUROC was 0.85 (95% CI, 0.78 to 0.91) (Suthanthiran et al., 2013). The diagnostic cutpoint in the CTOT-04 study that maximized sensitivity and specificity was -1.213, and TCMR was predicted with a sensitivity of 79% (95% CI, 67 to 91) and a specificity of 78% (95% CI, 71 to 84). In the current study in which centrifugation was not used to sediment urinary cells and the urinary cell lysate was prepared using ZFBP and the total RNA was isolated from the lysate using RNeasy Mini Kit, the three gene TCMR diagnostic signature score, computed using the CTOT-04 logistic regression equation, discriminated kidney allograft recipients with TCMR biopsies from those without TCMR biopsies and an AUROC of 0.83 (95% CI, 0.69 to 0.98) and TCMR was predicted with a sensitivity of 67% (95% CI, 35 to 89) and a specificity of 86% (95% CI, 67 to 95) using the CTOT-04 diagnostic threshold of -1.213. As reported in the results section, the TCMR diagnostic parameters - AUROC, sensitivity and specificity-were not different between the current WCHP-based study and the multicenter CTOT-04 study.

In our single center noninvasive diagnosis of BKVN validation study in which centrifugation was used to sediment the urine cell pellet and total RNA was isolated from the urinary cell lysate (Dadhania et al., 2010), ROC curve analysis yielded an AUROC of 0.99 (95% CI, 0.99 to 1.00, P<0.0001) and BKVN was diagnosed with a sensitivity of 100% (95% CI, 75.8 to 100) and a specificity of 97% (95% CI, 91 to 99). In the current study in which centrifugation was not used to sediment urinary cells and the urinary cell lysate was prepared using ZFBP and the total RNA was isolated from the lysate using the RNeasy Mini Kit, BKV VP1 mRNA copy number discriminated patients with BKVN biopsies from those with No Rejection with an AUROC of 1.000 (95% CI, 1.000 to 1.000, P<0.0001). In the current investigation, sensitivity was 86% (95% CI, 42 to 99) and the specificity was 100% (95% CI, 85 to 100) with the previously validated cutpoint of 6.5×10^8^ BKV VP1 mRNA copies per microgram of total RNA. As reported in the results section, the BKVN diagnostic parameters - AUROC, sensitivity and specificity-were not different between the current WCHP based study and our earlier BKVN validation study.

In the current investigation, we modified the ZFBP by the inclusion of a silica-membrane cartridge for mRNA enrichment and demonstrate noninvasive diagnosis of TCMR with performance characteristics similar to the CTOT-04 study, and noninvasive diagnosis of BKVN with performance characteristics similar to our BKVN validation study. Our development of WCHP for urinary cell mRNA profiling of kidney allograft recipients represent a significant advance towards not only portability of urinary cell mRNA profiling but also improved patient management by minimizing their visits to a laboratory, ambulatory clinic, or a hospital for the collection of urine, an excellent surrogate biospecimen for the invasively obtained kidney allograft biopsy (Verma et al., 2020).

## Data Availability

All data produced in the present study are available upon reasonable request to the authors.

## Acknowledgements

We thank Dr. Baogui Li and Dr. Joseph Schwartz for their original contributions to our urinary cell mRNA profiling studies. We thank our dedicated kidney transplant team at NYP-WCM and our patients - the true heroes of transplantation-in the iconic words of the pioneering transplant surgeon Professor Thomas E. Starzl.

## References

Abuhelaiqa E, Snopkowski C, Li C, Salvatore S, Lee JR, Muthukumar T, Lee JB, Hartono C, Ding R, Seshan SV, Suthanthiran M, Dadhania DM. Validation of a noninvasive prognostic signature for allograft failure following BK virus associated nephropathy. Clin Transplant. 2021 Feb;35(2):e14200.

Afaneh C, Muthukumar T, Lubetzky M, Ding R, Snopkowski C, Sharma VK, Seshan S, Dadhania D, Schwartz JE, Suthanthiran M: Urinary cell levels of mRNA for OX40, OX40L, PD-1, PD-L1, or PD-L2 and acute rejection of human renal allografts. Transplantation, 90: 1381–1387, 2010

Anglicheau D, Muthukumar T, Hummel A, Ding R, Sharma VK, Dadhania D, Seshan SV, Schwartz JE, Suthanthiran M: Discovery and validation of a molecular signature for the noninvasive diagnosis of human renal allograft fibrosis. Transplantation, 93: 1136–1146, 2012.

Bohl, D.L., and D.C. Brennan. 2007. BK virus nephropathy and kidney transplantation. Clin J Am Soc Nephrol 2 Suppl 1:S36–46.

Bradley MS, Boudreau MH, Grenier C, Huang Z, Murphy SK, Siddiqui NY: Urine RNA Processing in a Clinical Setting: Comparison of 3 Protocols. Female Pelvic Med Reconstr Surg, 25(3):247–251, 2019

Dadhania D, Muthukumar T, Ding R, Li B, Hartono C, Serur D, Seshan SV, Sharma VK, Kapur S, Suthanthiran M. Molecular signatures of urinary cells distinguish acute rejection of renal allografts from urinary tract infection. Transplantation. 2003 May 27;75(10):1752–4. doi: 10.1097/01.TP.0000063931.08861.56.

Dadhania, D., C. Snopkowski, R. Ding, T. Muthukumar, J. Lee, H. Bang, V.K. Sharma, S. Seshan, P. August, S. Kapur, and M. Suthanthiran. 2010. Validation of noninvasive diagnosis of BK virus nephropathy and identification of prognostic biomarkers. Transplantation 90:189–197.

Ding R, Medeiros M, Dadhania D, Muthukumar T, Kracker D, Kong JM, Epstein SR, Sharma VK, Seshan SV, Li B, Suthanthiran M: Noninvasive diagnosis of BK virus nephritis by measurement of messenger RNA for BK virus VP1 in urine. Transplantation, 74: 987–994, 2002

Ding R, Li B, Muthukumar T, Dadhania D, Medeiros M, Hartono C, Serur D, Seshan SV, Sharma VK, Kapur S, Suthanthiran M: CD103 mRNA levels in urinary cells predict acute rejection of renal allografts. Transplantation, 75: 1307–1312, 2003

Drachenberg, C.B., J.C. Papadimitriou, H.H. Hirsch, R. Wali, C. Crowder, J. Nogueira, C.B. Cangro, S. Mendley, A. Mian, and E. Ramos. 2004. Histological patterns of polyomavirus nephropathy: correlation with graft outcome and viral load. Am J Transplant 4:2082–2092.

Furness PN, Taub N; Convergence of European Renal Transplant Pathology Assessment Procedures (CERTPAP) Project. International variation in the interpretation of renal transplant biopsies: report of the CERTPAP Project. Kidney Int. 2001 Nov;60(5):1998–2012. doi: 10.1046/j.1523-1755.2001.00030.x. Erratum in: Kidney Int 2001 Dec;60(6):2429. PMID: 11703620.

Gomez-Alamillo C, Benito-Hernandez A, Ramos-Barron MA, Agueros C, Rodrigo E, Ruiz JC, Sanchez M, San Cosme L, Arias L: Analysis of Urinary Gene Expression of Epithelial-Mesenchymal Transition Markers in Kidney Transplant Recipients. Transplant Proc, 42(8):2886–8, 2010

Hart A, Smith JM, Skeans MA, Gustafson SK, Wilk AR, Castro S, Foutz J, Wainright JL, Snyder JJ, Kasiske BL, Israni AK: OPTN/SRTR 2018 Annual Data Report: Kidney. Am J Transplant, 20 Suppl s1: 20–130, 2020

Hirsch, H.H., P. Randhawa, and A.S.T.I.D.C.o. Practice. 2013. BK polyomavirus in solid organ transplantation. Am J Transplant 13 Suppl 4:179–188.

Li B, Hartono C, Ding R, Sharma VK, Ramaswamy R, Qian B, Serur D, Mouradian J, Schwartz JE, Suthanthiran M: Noninvasive diagnosis of renal-allograft rejection by measurement of messenger RNA for perforin and granzyme B in urine. N Engl J Med, 344: 947–954, 2001

Loupy, A., M. Haas, K. Solez, L. Racusen, D. Glotz, D. Seron, B.J. Nankivell, R.B. Colvin, M. Afrouzian, E. Akalin, N. Alachkar, S. Bagnasco, J.U. Becker, L. Cornell, C. Drachenberg, D. Dragun, H. de Kort, I.W. Gibson, E.S. Kraus, C. Lefaucheur, C. Legendre, H. Liapis, T. Muthukumar, V. Nickeleit, B. Orandi, W. Park, M. Rabant, P. Randhawa, E.F. Reed, C. Roufosse, S.V. Seshan, B. Sis, H.K. Singh, C. Schinstock, A. Tambur, A. Zeevi, and M. Mengel. 2017. The Banff 2015 Kidney Meeting Report: Current Challenges in Rejection Classification and Prospects for Adopting Molecular Pathology. Am J Transplant 17:28–41.

Lubetzky ML, Salinas T, Schwartz JE, Suthanthiran M. Urinary Cell mRNA Profiles Predictive of Human Kidney Allograft Status. Clin J Am Soc Nephrol. 2021 Oct;16(10):1565–1577. doi: 10.2215/CJN.14010820.

Mann RS, Carroll RB. Cross-reaction of BK virus large T antigen with monoclonal antibodies directed against SV40 large T antigen. Virology. 1984 Oct 30;138(2):379–85. doi: 10.1016/0042-6822(84)90365-9. PMID: 6093375

Morgan TA, Chandran S, Burger IM, Zhang CA, Goldstein RB. Complications of Ultrasound-Guided Renal Transplant Biopsies. Am J Transplant. 2016 Apr;16(4):1298–305. doi: 10.1111/ajt.13622.

Muthukumar T, Ding R, Dadhania D, Medeiros M, Li B, Sharma VK, Hartono C, Serur D, Seshan SV, Volk HD, Reinke P, Kapur S, Suthanthiran M: Serine proteinase inhibitor-9, an endogenous blocker of granzyme B/perforin lytic pathway, is hyperexpressed during acute rejection of renal allografts. Transplantation, 75: 1565–1570, 2003

Muthukumar T, Dadhania D, Ding R, Snopkowski C, Naqvi R, Lee JB, Hartono C, Li B, Sharma VK, Seshan SV, Kapur S, Hancock WW, Schwartz JE, Suthanthiran M. Messenger RNA for FOXP3 in the urine of renal-allograft recipients. N Engl J Med. 2005 Dec 1;353(22):2342–51. doi: 10.1056/NEJMoa051907.

Nickeleit V, Singh HK, Dadhania D, Cornea V, El-Husseini A, Castellanos A, Davis VG, Waid T, Seshan SV. The 2018 Banff Working Group classification of definitive polyomavirus nephropathy: A multicenter validation study in the modern era. Am J Transplant. 2021 Feb;21(2):669–680. doi: 10.1111/ajt.16189. Epub 2020 Aug 5.

Plattner BW, Chen P, Cross R, Leavitt MA, Killen PD, Heung M. Complications and adequacy of transplant kidney biopsies: A comparison of techniques. J Vasc Access. 2018 May;19(3):291–296. doi: 10.1177/1129729817747543.

Puri, K.S., K.R. Suresh, N.J. Gogtay, and U.M. Thatte. 2009. Declaration of Helsinki, 2008: implications for stakeholders in research. J Postgrad Med 55:131–134

Rampersad C, Balshaw R, Gibson IW, Ho J, Shaw J, Karpinski M, Goldberg A, Birk P, Rush DN, Nickerson PW, Wiebe C. The negative impact of T cell-mediated rejection on renal allograft survival in the modern era. Am J Transplant. 2022 Mar;22(3):761–771. doi: 10.1111/ajt.16883. Epub 2021 Nov 24. PMID: 34717048; PMCID: PMC9299170

Redfield RR, McCune KR, Rao A, Sadowski E, Hanson M, Kolterman AJ, Robbins J, Guite K, Mohamed M, Parajuli S, Mandelbrot DA, Astor BC, Djamali A. Nature, timing, and severity of complications from ultrasound-guided percutaneous renal transplant biopsy. Transpl Int. 2016 Feb;29(2):167–72. doi: 10.1111/tri.12660.

Røge R, Thorsen J, Tørring C, Ozbay A, Møller BK, Carstens J. Commonly used reference genes are actively regulated in in vitro stimulated lymphocytes. Scand J Immunol. 2007 Feb;65(2):202–9. doi: 10.1111/j.1365-3083.2006.01879.x. PMID: 17257226.

Snopkowski C, Salinas T, Li C, Stryjniak G, Ding R, Sharma V, Suthanthiran M. Urinary cell mRNA profiling of kidney allograft recipients: A systematic investigation of a filtration based protocol for the simplification of urine processing. J Immunol Methods. 2021 Nov;498:113132. doi: 10.1016/j.jim.2021.113132.

Suthanthiran M, Strom TB. Renal transplantation. N Engl J Med. 1994 Aug 11;331(6):365–76. doi: 10.1056/NEJM199408113310606.

Suthanthiran M, Schwartz JE, Ding R, Abecassis M, Dadhania D, Samstein B, Knechtle SJ, Friedewald J, Becker YT, Sharma VK, Williams NM, Chang CS, Hoang C, Muthukumar T, August P, Keslar KS, Fairchild RL, Hricik DE, Heeger PS, Han L, Liu J, Riggs M, Ikle DN, Bridges ND, Shaked A, Clinical Trials in Organ Transplantation 04 Study I: Urinary-cell mRNA profile and acute cellular rejection in kidney allografts. N Engl J Med, 369: 20–31, 2013

Tatapudi RR, Muthukumar T, Dadhania D, Ding R, Li B, Sharma VK, Lozada-Pastorio E, Seetharamu N, Hartono C, Serur D, Seshan SV, Kapur S, Hancock WW, Suthanthiran M: Noninvasive detection of renal allograft inflammation by measurements of mRNA for IP-10 and CXCR3 in urine. Kidney Int, 65: 2390–2397, 2004

Verma A, Muthukumar T, Yang H, Lubetzky M, Cassidy MF, Lee JR, Dadhania DM, Snopkowski C, Shankaranarayanan D, Salvatore SP, Sharma VK, Xiang JZ, De Vlaminck I, Seshan SV, Mueller FB, Suhre K, Elemento O, Suthanthiran M. Urinary cell transcriptomics and acute rejection in human kidney allografts. JCI Insight. 2020 Feb 27;5(4):e131552. doi: 10.1172/jci.insight.131552. PMID: 2102984; PMCID: PMC7101135.

Veronese FV, Manfro RC, Roman FR, Edelweiss MI, Rush DN, Dancea S, Goldberg J, Gonçalves LF. Reproducibility of the Banff classification in subclinical kidney transplant rejection. Clin Transplant. 2005 Aug;19(4):518–21. doi: 10.1111/j.1399-0012.2005.00377.x.

Wolfe RA, Ashby VB, Milford EL, Ojo AO, Ettenger RE, Agodoa LY, Held PJ, Port FK: Comparison of mortality in all patients on dialysis, patients on dialysis awaiting transplantation, and recipients of a first cadaveric transplant. N Engl J Med, 341: 1725–1730, 1999

